# Bringing conceptualizations of the health advocacy competence across the continuum of medical education together: a scoping review protocol

**DOI:** 10.1101/2024.12.09.24318699

**Authors:** Wilma RW Oosthoek, Dario Cecilio-Fernandes, Maarten FM Engel, Lars T van Prooijen, Suzie J Otto, Andrea M Woltman

## Abstract

**Introduction:** Health Advocacy (HA) is acknowledged as a core competence in medical education. However, varying and sometimes conflicting conceptualizations of HA exists, making it challenging to integrate the competence consistently. While this diversity highlight the need for a deeper understanding of HA conceptualizations, a comprehensive analysis across the continuum of medical education is absent in the literature. This protocol has been developed to clarify the conceptual dimensions of the HA competence in literature as applied to medical education.

**Methods and analysis:** The review will be conducted in line with the JBI methodology for scoping reviews. A comprehensive literature search was developed and already carried out in eight academic databases and Google Scholar, without restrictions on publication date, geography or language. Articles that describe the HA role among students and physicians that receive or provide medical education will be eligible for inclusion. Two independent reviewers will independently complete title and abstract screening, prior to full-text review of selected articles and data extraction on the final set. A descriptive-analytical approach will be applied for summarizing the data.

**Ethics and dissemination:** This scoping review does not involve human participants, as all evidence is sourced from publicly available databases. Therefore, ethical approval is not required for this study. The findings from this scoping review will be disseminated through submission to a high-quality peer-reviewed journal and presented at academic conferences. By clarifying the conceptualizations of HA, this review aims to contribute to a shared narrative that will strengthen the foundation for integrating the HA role into medical education.

**Trial registration number:** A preliminary version of this protocol was registered on the Open Science Framework (OSF) on 9 December 2024, and can be accessed at the following link: osf.io/6f94y.

**Article Summary:** 

**Strengths and limitations of this study:**

⇒ This review will maintain consistency and meet the quality standards established by other scoping reviews by adhering to the JBI methodology for scoping reviews and following to the Preferred Reporting Items for Systematic Reviews and Meta-Analyses extension checklist specifically designed for scoping reviews (PRISMA-ScR).
⇒ Two independent reviewers will conduct the title and abstract screening, full text screening and data extraction, including pilot exercises, to enhance the systematic rigor of the screening process and increase the reliability of the results.
⇒ The search strategy is systematically developed in collaboration with a medical librarian, who also serves as a co-author on this project.
⇒ This review ensures inclusivity by avoiding language restrictions, designing data visualizations for colorblind readers, and committing to open-access resources to align with FAIR (findable, accessible, interoperable and reusable) principles.
⇒ This review aims to pave the way toward a shared understanding of the health advocacy competence in medical education. However, while it contributes valuable insights to the discourse, it does not possess the capacity to inform policy or practice changes in the field.

## Introduction

Health Advocacy (HA) is a core competence for both current and future medical professionals to shape the healthcare system into one that is inclusive, accessible, affordable and sustainable for both individuals and populations (1,2). Although CanMEDS established the concept of HA in medical education (3), several other concepts related to HA have since emerged in the literature, making it challenging to integrate the competence consistently. Previous reviews on HA in medical education have addressed teaching and assessment methods related to HA at specific stages of medical education (4–8), but less attention has been given to the emerging variety of HA conceptualizations across the continuum of medical education. To bridge this gap, there is a need for a comprehensive analysis that brings clarity to the variety of HA conceptualizations in medical education.

CanMEDS defines HA competence as a role where physicians work with individuals, communities and populations to improve health outcomes, advocate for those they serve, and mobilize resources for change (9). However, this definition has been expanded, refined or challenged by others, incorporating various perspectives to address ambiguities around the role (10). HA is viewed as a clinical agent’s role, focusing on health promotion, compassionate patient-centered care and / or access to resources on an individual level (11– 14); as an activist role, addressing health injustices at both local and global levels (11– 13,15,16); as a personal role shaped by individual values and beliefs that drive perceptions and behaviors (17–19); and as a professional role influenced by the social contract between the medical profession and society, as well as by societal norms and values (18,20). Due to its diverse interpretations, overlap with other competencies, and fluid, unclear boundaries, the HA role is one of the least understood and assessed in medical education (10,21).

The wide range of conceptualizations allows medical students and physicians to align their advocacy efforts with their personal values, providing them with a sense of purpose and fulfillment (17,22). While the flexibility of the role offers opportunities for change, it also generates ethical tensions. The conceptual boundaries between the health advocacy role and medical expert role as defined and described by CanMeds are often blurred, contributing to conflicting views on how far medical professionals should go in promoting societal or systemic reform as part of their professional duties (23,24). Medical professionals could feel uncertain about how to balance these roles (12,16,25), a problem known as “dual agency” (26). Along with that, HA conceptualizations have shifted over time from a collective profession-wide responsibility to an individual physician’s role (27), with recent perspectives advocating for a focus on planetary health (28). This shift raises questions about which aspects of HA should be addressed at a profession-wide level, which fall under the responsibility of an individual physician, and which are the responsibility of society and its citizens (20,29).

The evolving diversity in HA conceptualizations highlights the need for an in-depth understanding of the HA role within medical education. A clearer understanding of HA conceptualizations will facilitate a shared narrative that fosters the foundation of an educational environment where HA competencies can reach their full potential. Currently, a comprehensive analysis of HA role conceptualizations across the continuum of medical education is lacking in literature. To address this gap, the primary purpose of this scoping review is to clarify conceptual dimensions of the HA competence in literature as applied to medical education. In this paper we used the term “health advocacy” to refer to all of the conceptual dimensions that compose it.

### Review questions

What is known about the concept of HA competence in medical education? This question is broken down into sub-questions:

- What is the nature and extent of the scientific literature on the HA role in medical education?
- What are the conceptual dimensions and boundaries of the HA role in medical education?
- How do these conceptualizations differ over each phase of medical education?
- How do these conceptualizations differ over time?

We decided on a scoping review since it is well-suited for bringing together and reporting upon heterogeneous conceptualizations in literature (30). A preliminary search of Embase, Medline, Cochrane and Google Scholar was conducted. No comprehensive review of HA conceptualizations over the whole scope of medical education was identified. By viewing the perspectives side by side, this review will lay the groundwork for future studies, whether through a consensus-building process toward a shared understanding of HA in medical education or a focused analysis of HA in specific education contexts or settings. We further propose that identifying points of convergence, where agreement on key elements of HA is achieved, will help establish a shared understanding of the role. At the same time, recognizing areas of divergence, where agreement may be challenging or impossible in nature, will inform future discussions regarding what should be taught and assessed concerning HA competencies.

## Methods and analysis

### Protocol and registration

The proposed scoping review will follow the JBI (formerly Joanna Briggs Institute) methodology for conducting scoping reviews and adhere to the Preferred Reporting Items for Systematic Reviews and Meta-Analyses extension for Scoping Reviews (PRISMA-ScR) checklist (31). Additionally, the best practice guidance and reporting items for the development of scoping review protocols (32) and recommendations for extraction, analysis and presentation of results in scoping reviews (33) will be used. Our protocol serves as a blueprint for the scoping review and is intended to reduce potential reporting biases. Any changes made to the protocol during the review process will be clearly documented and explained in the scoping review. The protocol was preregistered with the Open Science Framework (OSF).

### Eligibility criteria

#### Population

For this review, only articles that focus on students and physicians that receive or provide medical education, e.g. medical students / trainees, residents, fellows, and practicing physicians across all levels of experience, will be eligible. This encompass physicians in both general practice and specialized medical fields, as well as those engaged in formal educational roles, such as clinical educators. Additionally, articles addressing the medical profession as a whole will also be considered eligible.

#### Concept

The concept under investigation in this scoping review is “health advocacy competence”, as defined by the CanMEDS framework (9). We carefully selected terms for “health advocacy” and “competence”. Eligible terms for HA will include “advocacy,” “activism,” and “agency” related to “health” if they are the primary study topic or key component of theoretical discussions on competence. However, broader terms like “involvement” “engagement”, “responsibility”, “accountability”, “health promotion”, “system change” and “health equity” will not be eligible unless the authors explicitly link such terms to HA. We predetermined that we will not infer it on the authors’ behalf. Eligible terms for “competence” will include all activities that relate to attainment, retention and/or loss of knowledge, skills and attitudes, such as “life-long learning” and “skill”.

### Context

This review focuses on HA in the medical education context, as defined following the definition of the World Medical Association (2020): “Medical education consists of basic medical education, postgraduate medical education, and continuing professional development. Medical education is a dynamic process that commences at the start of basic medical education (medical school) and continues until a physician retires from active practice (34).” The different phases of medical education, as undergraduate, graduate, postgraduate and life-long / continuous learning, will be eligible. Additionally, articles addressing the medical curriculum as a whole will be considered eligible. We excluded the term “foundation” due to its multiple meaning beyond the UK-specific postgraduate “F1 and F2” years. No restrictions were placed on publication date, geography or language, as we aim for a comprehensive understanding of HA conceptualizations over borders of countries, cultures, traditions and healthcare systems.

### Types of information sources

Eligible sources include theoretical (e.g. reviews) and empirical studies (qualitative, quantitative or mixed method), as well as opinion-based articles as perspectives and editorials. Conference abstracts and notes / comments were excluded to manage the overall number of results. Additionally, most conference abstracts are eventually published in full within the medical education and practice literature, providing further justification for their exclusion.

### Search strategy

The search strategy was developed through a collaborative effort between an experienced information specialist (M.E.) and the lead author (W.O.), and revised by the other team members. The search was developed in Embase.com, optimized for sensitivity and then translated to other databases following the method as described by Bramer et al. (2018) (35). The search contained terms for 1) “health advocacy”, 2) “competence” and 3) “medical education”. Relevant terms from titles, abstracts, and index terms were explored and refined through discussions between the research team, and once agreed upon, were incorporated into the search. Terms were combined with Boolean operators (AND, OR), and proximity operators were used to combine terms into phrases. The search was carried out in eight electronic databases on October 29^th^, 2024: Embase, Medline, Cochrane, Web of Science, CINAHL, ERIC, PsycInfo and LILACS. Additionally, a search was performed in Google Scholar from which the 100 highest-ranked references were downloaded using the software Publish or Perish v8.0. The full search strategies of all databases are available in Supplemental Material 1, and will be updated prior to the submission of the scoping review to ensure all data is current using the methods as described by Bramer et al (2017) (36). The search strategies for Medline and Embase used relevant thesaurus terms from Medical Subject Headings (MeSH) and Emtree respectively. In all databases, terms were searched in titles, abstracts and author keywords. The searches in Embase, Medline, Web of Science, ERIC, Cinahl and PsycInfo were limited to exclude conference abstracts and notes/comments. We did not browse unindexed journals in the field. The references were imported into EndNote and duplicates were removed by M.E. using the method as described by Bramer et al. (2016) (37). Additionally, Covidence identified duplicates that have been inadvertently missed.

### Data management

The web-based collaboration software platform Covidence (2025) will be used to screen articles and track citations to search for additional articles. The Erasmus MC research project management system PANAMA will be used for study registration and archiving. Zotero v7.0. is used to manage references.

### Selection process

Two reviewers (W.O. and L.P.) will independently apply the inclusion criteria for title and abstract screening. To facilitate calibration, the first meeting will focus on creating a shared understanding of the inclusion criteria. A pilot test on the first hundred articles will be performed and repeated till 90% of agreement is reached. Title and abstract screening will be followed by full-text reading of the included articles. If an article does not match the inclusion criteria after full-text reading, the article will be excluded if both reviewers agreed. If disagreements arise or if adjustments to the iteratively developed exclusion criteria are needed during the different stages of the screening process, a third and fourth team member (D.C.F. and S.O.) will be consulted. For articles written in other languages than could be translated by our team (e.g. Dutch, English, Spanish and Portuguese), online translation tools such as Erudite (Erasmus University’s internal AI chatbot, 2025) or DeepL (2025) will we used. If the translations from these tools are unclear, the article will be forwarded to a colleague who is a native speaker of the original language. Any changes to inclusion / exclusion criteria or steps taken to resolve inconsistencies will be thoroughly documented. Excluded sources with reason for exclusion will be reported for full-text level and included in the appendices of the scoping review. As last step the reference lists of retrieved non-included relevant review articles and of the included references, as well as articles citing these papers, will be scanned for relevant references missed by the search using the methods as described by Bramer et al (2018) (38). The selection process will be presented in both a narrative and PRISMA flow diagram format (see Supplemental Material 2) (39).

### Data extraction

To answer the research questions, two researchers will extract the following: Digital Object Identifier (DOI); authors; e-mail address of corresponding author; year of publication; country of origin; funding source; title of publication; content/topic(s); study objective(s)/aim(s); type of evidence source; study design; study population; language of data; main outcome; conclusion; future directions; conceptualizations; and practical applications. Our study pre-defined various forms of HA conceptualizations in medical education, including definitions, frameworks, models, theories and conceptual metaphors (for detailed information see data guidance sheet, Supplemental Material 3). Practical applications stem from underlying conceptualizations and serve as their reflections, but they are distinct from conceptualizations themselves. Therefore, we extract practical applications (e.g. examples that demonstrate HA in action) separately. A data charting template, based on the guidance sheet, was developed to support the authors systematically extract and compare the results. To minimize researcher bias and ensure the reliability and feasibility of the template, a pilot test will be conducted by two members of the team (W.O. and L.P.), with iterations made until 90% agreement is reached. Any discrepancies will be addressed during regular meetings with the third and fourth team member (D.C.F. and S.O.). Following the pilot phase, data extraction will proceed sequentially by each reviewer, with regular checks for consistency. The data extraction process will be iterative, allowing for updates to the guidance sheet and charting form as the study progresses. If necessary, authors of the original studies may be contacted to request missing or supplementary data. Any deviations from the protocol will be carefully documented in the scoping review.

### Data analysis and presentation

The scoping review will be written in English. A descriptive-analytical approach will be used for summarizing the data. Quantitative data will include a descriptive numerical summary of the study characteristics and frequency counts of data extraction items. A basic qualitative content analysis will be conducted using inductive open coding to group the different HA conceptualizations into overarching categories, following Pollock et al. (2023) (33). Then, the range of general codes will be presented over time, categorized into four periods: <1996 (period before the year in which HA was formally defined within the CanMEDS framework in 1996 (3)), 1996 – 2010, 2010 – 2020, 2020 – current. These timeframes may be adjusted as we analyze the data, allowing the findings to guide our final decisions. Subsequently, the range of general codes will be presented over each phase of medical education, using four pillars: preclinical, clinical, postgraduate and continual professional development. Data will be presented visually in figures whenever possible, with consideration to color blindness using Color Oracle v1.3.

## Data Availability

All data produced in the present study are available upon reasonable request to the authors

## Summary

This scoping review aims to establish a foundational understanding of how health advocacy is conceptualized across the spectrum of medical education. By exploring the diverse conceptualizations of HA, the review seeks to clarify the varying interpretations of HA roles within the medical education field. The findings will not only provide a basis for future research, but also drive meaningful change by clarifying where consensus can be built and what constitutes the core of HA competencies. Recognizing both areas of convergence and divergence will guide discussions on how HA should be taught, assessed and implemented in the whole continuum of medical education.

## Ethics and dissemination

This scoping review does not involve any human participants. Consequently, there is no need for ethical approval. The dissemination of the findings will take place through the submission of a single manuscript to a high-quality peer-reviewed journal and will be disseminated through scientific literature and presentations at key conferences.

## Acknowledgements

The authors would like to express their gratitude to Dr. E.F. van Beeck, Education Lead of the Erasmus MC Public Health Department, for his guidance in shaping the initial direction and purpose of this protocol.

## Author contributions

W.O. drafted the protocol and reviewed all sections in iterative sessions with S.O., D.C.F. and A.W. M.E. collaborated closely with the research team in developing the search strategy and reviewed the method section of the protocol. L.P. is a student contributor who critically reviewed protocol and gave suggestions on aims, relevance and scope of the search strategy to the research team. W.O. and L.P. piloted the screening criteria and revised them in one interactive meeting with S.O. and D.C.F. The pre-final protocol was critically assessed with the entire research team. All authors read and approved the study protocol.

## Funding

No external grants were received for this project from any funding agency, whether in the public, commercial, or non-profit sectors.

## Patient and public involvement

This work analyses existing articles, and therefore involves no patients or citizens.

## Competing interests

The authors have no competing interests to declare.

## Declaration of generative AI and AI-assisted technologies in the writing process

During the preparation of this work the author(s) used Erasmus University’s (EUR) internal AI chatbot Erudite for translation purposes and language refinement. Erudite is available for users as a chat in Microsoft Teams, o□ered within the existing agreements the EUR has with Microsoft. After using this tool, the author(s) reviewed and edited the content as needed and take(s) full responsibility for the content of the published protocol.

## Notes

### Competing Interest Statement

The authors have declared no competing interest.

### Funding Statement

This study did not receive any funding

