## Supplemental Material 1 for "Bringing conceptualizations of the health advocacy competence across the continuum of medical education together: a scoping review protocol"

**Supplemental material 1 Search Strategy**

Searched November 26^th^, 2024

| **Database searched** | **Platform** | **Years of coverage** | **Records** | **Records after duplicates removed** |
| --- | --- | --- | --- | --- |
| Embase | Embase.com | 1971 - Present | 1686 | 422 |
| Medline ALL | Ovid | 1946 - Present | 1541 | 1532 |
| Cochrane Central Register of Controlled Trials | Wiley | 1992 - Present | 166 | 131 |
| Web of Science Core Collection* | Web of Knowledge | 1975 - Present | 1665 | 994 |
| CINAHL Plus** | EBSCO | 1982 - Present | 1031 | 436 |
| ERIC | Ovid | 1965 - Present | 1361 | 1275 |
| PsycINFO | Ovid | 1806 - Present | 969 | 537 |
| LILACS | https://lilacs.bvsalud.org/en/ |  | 3 | 1 |
| Additional Search Engines: Google Scholar*** (100 top ranked) | | | 100 | 16 |
| **Total** | | | **8522** | **5344** |

*Science Citation Index Expanded (1975-present) ; Social Sciences Citation Index (1975-present) ; Arts & Humanities Citation Index (1975-present) ; Conference Proceedings Citation Index- Science (1990-present) ; Conference Proceedings Citation Index- Social Science & Humanities (1990-present) ; Emerging Sources Citation Index (2005-present)

*Exact search turned on in Web of Science Core Collection

**Limited to Academic Journals

***Google Scholar was searched via "Publish or Perish" to download the results in EndNote.

No other database limits were used than those specified in the search strategies

*Excluded publication types were conference abstracts, Notes and for Cochrane only: trail registries*

**Embase 1686**

('health advocacy'/de OR 'health activism'/de OR activism/de OR 'consumer advocacy'/de OR (((health OR clinical) NEAR/3 (advocacy OR advocate)) OR ((clinical OR health) NEXT/1 agency) OR activism*):ab,ti,kw OR (advocacy OR activism OR advocate):ti) AND (education/exp OR skill/de OR aptitude/de OR learning/de OR 'lifelong learning'/de OR competence/de OR 'professional competence'/de OR (educat* OR life-long-learn* OR skill* OR aptitude* OR training* OR schooling OR learning OR tutoring OR coaching OR lecturing OR lesson* OR apprenticeship* OR internship* OR workshop* OR competen*):ab,ti,kw) AND (clinician/de OR 'hospital physician'/exp OR 'medical student'/exp OR resident/de OR 'general practitioner'/de OR physician/exp OR 'medical profession'/de OR (clinician* OR physician* OR general-practitioner* OR ((medical) NEAR/3 (student* OR undergraduate OR graduate OR postgraduate OR trainee* OR campus OR school)) OR ((medical) NEAR/3 (profession* OR curriculum)) OR resident* OR house-officer* OR surgeon* OR psychiatrist* OR pediatrician* OR paediatrician* OR fellow OR fellows):ab,ti,kw OR student*:ti) NOT ([Conference Abstract]/lim OR [Conference Review]/lim) NOT (note/de)

**Medline 1541**

(Political Activism/ OR Consumer Advocacy/ OR (((health OR clinical) ADJ3 (advocacy OR advocate)) OR ((clinical OR health) ADJ agency) OR activism*).ab,ti,kf. OR (advocacy OR activism OR advocate).ti.) AND (exp Education/ OR Aptitude/ OR Learning/ OR Professional Competence/ OR (educat* OR life-long-learn* OR skill* OR aptitude* OR training* OR schooling OR learning OR tutoring OR coaching OR lecturing OR lesson* OR apprenticeship* OR internship* OR workshop* OR competen*).ab,ti,kf.) AND (exp Physicians/ OR Students, Medical/ OR General Practitioners/ OR Health Occupations/ OR (clinician* OR physician* OR general-practitioner* OR ((medical) ADJ3 (student* OR undergraduate OR graduate OR postgraduate OR trainee* OR campus OR school)) OR ((medical) ADJ3 (profession* OR curriculum)) OR resident* OR house-officer* OR surgeon* OR psychiatrist* OR pediatrician* OR paediatrician* OR fellow OR fellows).ab,ti,kf. OR student*.ti.) NOT (congres* OR abstract*).pt. NOT (Comment.pt.)

**Cochrane 166**

((((health OR clinical) NEAR/3 (advocacy OR advocate)) OR ((clinical OR health) NEXT/1 agency) OR activism*):ab,ti,kw OR (advocacy OR activism OR advocate):ti) AND ((educat* OR life-long-learn* OR skill* OR aptitude* OR training* OR schooling OR learning OR tutoring OR coaching OR lecturing OR lesson* OR apprenticeship* OR internship* OR workshop* OR competen*):ab,ti,kw) AND ((clinician* OR physician* OR general-practitioner* OR ((medical) NEAR/3 (student* OR undergraduate OR graduate OR postgraduate OR trainee* OR campus OR school)) OR ((medical) NEAR/3 (profession* OR curriculum)) OR resident* OR house-officer* OR surgeon* OR psychiatrist* OR pediatrician* OR paediatrician* OR fellow OR fellows):ab,ti,kw OR student*:ti) NOT ("conference abstract":kw OR Trial registry record:pt)

**Web of Science 1665**

TS=(((((health OR clinical) NEAR/2 (advocacy OR advocate)) OR ((clinical OR health) NEAR/1 agency) OR activism*)) AND ((educat* OR life-long-learn* OR skill* OR aptitude* OR training* OR schooling OR learning OR tutoring OR coaching OR lecturing OR lesson* OR apprenticeship* OR internship* OR workshop* OR competen*)) AND ((clinician* OR physician* OR general-practitioner* OR ((medical) NEAR/2 (student* OR undergraduate OR graduate OR postgraduate OR trainee* OR campus OR school)) OR ((medical) NEAR/2 (profession* OR curriculum)) OR resident* OR house-officer* OR surgeon* OR psychiatrist* OR pediatrician* OR paediatrician* OR fellow OR fellows))) NOT DT=(Meeting Abstract OR Meeting Summary) NOT DT=(Note)

**CINAHL 1031**

(MH Consumer Advocacy OR AB(((health OR clinical) N2 (advocacy OR advocate)) OR ((clinical OR health) N1 agency) OR activism*) OR TI(((health OR clinical) N2 (advocacy OR advocate)) OR ((clinical OR health) N1 agency))) AND (MH Education+ OR MH Aptitude+ OR MH Learning OR AB(educat* OR life-long-learn* OR skill* OR aptitude* OR training* OR schooling OR learning OR tutoring OR coaching OR lecturing OR lesson* OR apprenticeship* OR internship* OR workshop* OR competen*) OR TI(educat* OR life-long-learn* OR skill* OR aptitude* OR training* OR schooling OR learning OR tutoring OR coaching OR lecturing OR lesson* OR apprenticeship* OR internship* OR workshop* OR competen*)) AND (MH Physicians+ OR MH Students, Medical+ OR MH Physicians, Family OR MH Health Occupations OR AB(clinician* OR physician* OR general-practitioner* OR ((medical) N2 (student* OR undergraduate OR graduate OR postgraduate OR trainee* OR campus OR school)) OR ((medical) N2 (profession* OR curriculum)) OR resident* OR house-officer* OR surgeon* OR psychiatrist* OR pediatrician* OR paediatrician* OR fellow OR fellows) OR TI(clinician* OR physician* OR general-practitioner* OR ((medical) N2 (student* OR undergraduate OR graduate OR postgraduate OR trainee* OR campus OR school)) OR ((medical) N2 (profession* OR curriculum)) OR resident* OR house-officer* OR surgeon* OR psychiatrist* ORpediatrician* OR paediatrician* OR fellow OR fellows OR student*)) NOT (MH Abstracts)AND PT academic*

**ERIC 1361**

(Activism/ OR (((health OR clinical) ADJ3 (advocacy OR advocate)) OR ((clinical OR health) ADJ agency) OR activism*).ab,ti. OR (advocacy OR activism OR advocate).ti.) AND (exp Education/ OR Aptitude/ OR Learning/ OR Lifelong Learning/ OR Competence/ OR (educat* OR life-long-learn* OR skill* OR aptitude* OR training* OR schooling OR learning OR tutoring OR coaching OR lecturing OR lesson* OR apprenticeship* OR internship* OR workshop* OR competen*).ab,ti.) AND (exp Physicians/ OR Medical Students/ OR Health Occupations/ OR (clinician* OR physician* OR general-practitioner* OR ((medical) ADJ3 (student* OR undergraduate OR graduate OR postgraduate OR trainee* OR campus OR school)) OR ((medical) ADJ3 (profession* OR curriculum)) OR resident* OR house-officer* OR surgeon* OR psychiatrist* OR pediatrician* OR paediatrician* OR fellow OR fellows).ab,ti. OR student*.ti.) NOT (congres* OR abstract*).pt. NOT (Comment.pt.)

**PsycINFO 969**

(Activism/ OR (((health OR clinical) ADJ3 (advocacy OR advocate)) OR ((clinical OR health) ADJ agency) OR activism*).ab,ti. OR (advocacy OR activism OR advocate).ti.) AND (exp Education/ OR Learning/ OR exp Competence/ OR (educat* OR life-long-learn* OR skill* OR aptitude* OR training* OR schooling OR learning OR tutoring OR coaching OR lecturing OR lesson* OR apprenticeship* OR internship* OR workshop* OR competen*).ab,ti.) AND (exp Physicians/ OR Medical Students/ OR General Practitioners/ OR (clinician* OR physician* OR general-practitioner* OR ((medical) ADJ3 (student* OR undergraduate OR graduate OR postgraduate OR trainee* OR campus OR school)) OR ((medical) ADJ3 (profession* OR curriculum)) OR resident* OR house-officer* OR surgeon* OR psychiatrist* OR pediatrician* OR paediatrician* OR fellow OR fellows).ab,ti. OR student*.ti.) NOT (congres* OR abstract*).pt. NOT (Comment.pt.)

**LILACS 3**

(“health advocacy” OR activism) AND (education OR “life long learning” OR skill OR skills OR aptitude OR training OR schooling OR learning OR tutoring OR coaching OR lecturing OR lesson OR apprenticeship OR internship OR workshop OR competen*) AND (clinician OR physician OR “general practitioner” OR ”medical student” OR “medical undergraduate” OR “medical graduate” OR “medical postgraduate” OR “medical trainee” OR ”medical profession” OR resident OR “house officer” OR surgeon OR psychiatrist OR pediatrician OR paediatricianOR fellow OR fellows)

**Google Scholar 100**

‘health advocacy’ competence|education|learning|skill|training clinician|physician|’general practitioner’|’medical student|graduate|undergraduate|trainee|profession’|resident|surgeon
