## Supplemental Material 2 for "Bringing conceptualizations of the health advocacy competence across the continuum of medical education together: a scoping review protocol"

**Supplemental material 2 PRISMA Flow Chart**

**Identification of studies via databases and registers**

Records removed *before screening*:

Duplicate records removed (n = 3178)

Records marked as ineligible by Covidence (n = 1)

Records removed for other reasons (n = ?)

Records identified from:

Databases (n = 8522)

EMBASE (n = 1686)

Medline (n = 1541)

Cochrane (n = 166)

Web of Science (n = 1665)

CINAHL (n = 1031)

ERIC (n = 1361)

PsychINFO (n = 969)

LILACS (n = 3)

Google Scholar (n=100)

**Identification**

Reports sought for retrieval

(n = )

Reports not retrieved

(n = )

Records excluded

(n = )

Records screened

(n ≤ 5343)

**Screening**

Reports excluded:

Reason 1 (n = )

Reason 2 (n = )

Reason 3 (n = )

etc.

Reports assessed for eligibility

(n = )

Studies included in review

(n = )

Reports of included studies

(n = )

**Included**
