## Supplemental Material 3 for "Bringing conceptualizations of the health advocacy competence across the continuum of medical education together: a scoping review protocol"

**Supplemental material 3 Data guidance sheet**

| **Data type** | **Explanation** |
| --- | --- |
| *Basic characteristics* | |
| Digital Object identifiers (DOI) | The full DOI listed in the metadata of the article. |
| Authors | The surname of authors. For more than two authors *et al.* If there was an organization as author, the whole name of organization. |
| Email address of corresponding author | Email dress as mentioned on the article. |
| Year of publication | The year the article was published. |
| Country of origin | Country where study is conducted. If no data was collected, country of first and last author. |
| Funding source | Funding source as mentioned in de “funding source” section of article. |
| *General content* | |
| Title of publication | Title as mentioned on top of the article. |
| Content topic(s) | Type of HA competency discussed as represented in title, abstract or keywords. |
| Study objective(s)/aim(s) | Study objective(s)/aims as mentioned at the end of introduction or start of method section. |
| Type of evidence source | Primary research: peer-reviewed research articles  Epidemiology: articles that have used population-level datasets  Evidence syntheses: e.g. narrative reviews, systemic reviews, scoping reviews, mapping reviews  Innovation reports  Discussion articles: e.g. essays, perspectives  Editorials |
| Study design | Qualitative, Quantitative, Mixed, Reviews, Others and No design (e.g. perspective, essay). |
| Study population | E.g. pre-clinical / clinical / postgraduate students (e.g. residents, fellows), and physicians in different specialties and phases of career. If applicable, add information on the sample size used for the outcome. |
| Language of data | Language of data, state if language of abstract is different than rest of article (state both in such a case) |
| Main outcome | Primary outcome measure(s) identified in the study. The main outcome is not explicitly mentioned, extract the primary objective of the study and any related measures (e.g., advocacy behavior change). Indicate the units of measurement and time points assessed. Note any missing or ambiguous data and flag for clarification with the study authors if needed. |
| Conclusion | Answer to the research question (no more than three sentences) |
| Future directions | Most important future directions mentioned by the authors (no more than three sentences) |
| *In depth content* | |
| Conceptualizations (e.g. definitions, frameworks, models, theories, interpretations and / or conceptual metaphors) | Statements that define or describe the concept of “health advocacy” or related terms, including references if the authors cite specific definitions. Additionally, extract any frameworks, models, theories, interpretations and /or metaphors that the authors use or refer to in structuring or explaining health advocacy within the context of medical education, including references if the authors cite specific frameworks, models or theories.  For interpretations, only subjective insights or viewpoints of the authors regarding health advocacy’s importance, role or impact should be extracted. For conceptual metaphors, only the “source conceptual domain” (e.g. “ Robin Hood”) and “target conceptual domain”(e.g. Health Advocacy role”) as mentioned by Lakoff & Johnson (40), and the meaningful context in which the metaphor is embedded by the authors should be extracted. Only interpretations and metaphors by the authors themselves should be extracted. No participant quotes from the results section should be extracted. |
| Practical applications | Examples or cases that show how health advocacy is practiced and applied in medical education, mentioned by authors in the article (e.g. teaching activity, implementation tool, assessment instrument) |
| *Other* | |
| Relevant notes of data extractor |  |
| Relevant illustrations |  |
